# Level of partograph utilization and its predictors among obstetrics care givers in Ilu Aba Bor Zone, Southwest Ethiopia: a cross-sectional study

**DOI:** 10.1101/2023.01.20.23284834

**Authors:** Getamesay Aynalem Tesfaye, Fentaneh Teshome Chanie, Ebissa Negera Gemechu

**Affiliations:** Public Health Department, College of Medicine and Health Sciences, Jinka University, P.O. Box 174, 4420 Jinka, Ethiopia; Chagni Health Center, Chagni, Ethiopia; Public Health Department, Faculty of Medicine and Health Sciences, Mettu University, Mettu, Ethiopia

**Keywords:** Ethiopia, Ilu Aba Bor, maternal death, obstetrics care, partograph utilization

## Abstract

**Background:** Worldwide, several thousands of mothers die per a year due to pregnancy and birth related complications. Most of maternal deaths can be minimized using partograph routinely. In spite of the priceless importance of partograph in reducing maternal death, its level of utilization and associated factors has been little known among obstetrics care providers in Ilu Aba Bor Zone.

**Objective:** This study aimed at assessing the level of partograph utilization and associated factors among obstetric care givers in Ilu Aba Bor Zone, South West Ethiopia.

**Methods:** An institution-based cross-sectional study design was employed. A structured self-administered and pretested questionnaire adapted from available literatures were used. In addition to descriptive statistics, logistic regression analysis was applied to assess association.

**Results:** The level of partograph utilization among obstetrics care providers in the study area was 32.8%. Receiving on-job training on partograph (AOR (Adjusted Odds Ratio) = 2.21, 95%CI (Confidence Interval) = 1.19, 4.11), working in a hospital compared to working in a health center (AOR = 2.43, 95%CI = 1.01, 5.82), having BSc (Bachelor of Science) and above educational status in contrast to having Diploma (AOR = 3.12, 95%CI = 1.59, 6.12), and having partograph in a health facility (AOR= 4.19, 95%CI = 2.12, 8.29) were positively associated with partograph use.

**Conclusion:** Partograph utilization level was much lower than World Health Organization recommendation. On-job training on partograph, work place, educational status, and partograph availability were predictors of level of partograph utilization among the obstetric care givers.

## Introduction

Globally, in 2015, the annual number of maternal deaths was 303,000, while the approximate global lifetime risk of a maternal death was 1 in 180.^1^ In the same year, approximately 99% (302,000) of the global maternal deaths occurred in developing countries, with sub-Saharan Africa alone accounting for roughly 66% (201,000). The 2016 Ethiopia Demographic and Health Survey estimated that maternal mortality ratio in Ethiopia is 412 deaths per 100,000 live births.^2^

Most of maternal deaths are the direct result of complications arising during pregnancy, delivery, or the puerperium that includes hemorrhage, hypertensive disorders of pregnancy, sepsis, prolonged labor, and unsafe abortion.^3,4^ If a woman with prolonged or obstructed labor does not get timely and effective management, she may die of uterine rupture or infection.

Partograph is one of the simplest tools employed to prevent maternal death by helping obstetrics health care providers identify slow progress in labor early, and initiate appropriate interventions to prevent prolonged and obstructed labor.^5,6^ It can be highly effective in reducing complications from prolonged labor for the mother and for the new-born.^7-9^

Despite the invaluable and affordable significance of partograph in reducing maternal death, its utilization remained near to the ground in low- and middle-income countries.^10^ A study conducted in Kenya revealed that most of nurses (55.5%) did not duly complete partograph, while elsewhere in South Africa, only 54 (79.4%) routinely used partograph.^11,12^ Similarly, studies in other parts of Africa showed that the use of partograph by health care providers is below World Health Organization (WHO) recommendation for its routine usage.^13-15^ Even though Ethiopia has set the Sustainable Development Goal target of reducing maternal mortality to 199/100,000 in 2030,^16^ the use of partograph to prevent maternal death is still low in various parts of the country.^17-19^

Different factors could hinder routine and proper utilization of partograph. For instance, a systematic review conducted in 2014 revealed that professional skills, clinical leadership and quality assurance, and the organizational environment within the wider provision of obstetric care were barriers to partograph use.^10^ According to a cross sectional study in Nigeria, factors affecting utilization of partograph were little or no knowledge of the partograph, non-availability, shortage of staff, and the fact that it is time-consuming to use.^20^ Getting on job training, being knowledgeable on partograph and having favorable attitude towards partograph were positively associated with partograph utilization in a study conducted in Central Ethiopia.^21^

Little has been known about partograph utilization and its determinants among obstetrics care providers working in public health institutions found in Ilu Aba Bor Zone, South West Ethiopia. Therefore, this study aimed at assessing the level of partograph utilization and associated factors among obstetric care givers in the study area. Eventually, this study will be substantial to reduce maternal mortality by encouraging partograph utilization among obstetrics care givers through tackling factors that hinders partograph utilization. This study will also provide a base line information for managers and researchers.

## Methods and Materials

### Study Area

The study was conducted at selected public health institutions that are found in Ilu Aba Bor Zone, South West Ethiopia. The capital city of the Ilu Aba Bor Zone is Mettu, which is located 600 kilometers away south west of Addis Ababa. Ilu Aba Bor Zone lies at attitude of 1,500-2,000 meters above sea level. According to the Health Department of Ilu Aba Bor Zone, it has 2 hospitals, 41 health centers and 273 health posts. There are 840 health professionals working in these public health institutions, of which 590 are obstetrics c are givers.

### Study Design and Period

An institution-based cross-sectional study was carried out from July 1 to August 30, 2020.

### Population

The source population of this study are all obstetrics care givers working in public health institutions of Ilu Aba Bor Zone. Whereas, the study population of this study are those selected obstetrics care givers working in randomly selected public health institution of Ilu Aba Bor Zone.

### Eligibility Criteria

Obstetrics care givers who were working in labor and delivery ward in regular and duty time were included in this study. On the other hand, those obstetric care givers who were sick, and who went away for a long-time leave were excluded from this study.

### Sample Size Determination

The sample size of the study was determined by using a single proportion formula based on a 40.2% level of partograph utilization in Central Ethiopia,^21^ 95% confidence interval, and margin of error 0.05. Hence, the formula yielded a preliminary sample size of 369. Since the source population size is below 10,000, using correction formula, the sample size became 227. Finally, after considering 10% non-response rate, the final sample size became 250.

### Sampling Technique

Out of 2 hospitals and 41 health centers which were providing public obstetrics service in Ilu Aba Bor Zone, 27 health facilities (26 health centers and 1 hospital) were selected by simple random technique to meet the sample size after stratifying them to the level of service they give. Then, after allocating the sample size proportionally to each health facility, obstetric caregivers were selected randomly using lottery method.

### Operational Definitions

#### Public health institution

Health centers and hospitals operated and controlled by government.

#### Obstetric caregivers

Health professionals including medical doctors, emergency and obstetrics surgeons, midwiferies, nurses, and health officers who are responsible for giving labor follow up and delivery service.^22^

#### Partograph utilization

When an obstetric caregiver uses partograph routinely for all laboring mother.^22^

### Data Collection Procedure and Instruments

A structured self-administered questionnaire adapted from available literatures were used.^23-25^ The questionnaire was consisted of questions about socio-demographic and professional characteristics of participants, and also about the outcome variable which is utilization of partograph. Data collection were carried out by three Health Officers from a non-selected institution, and the data collectors were supervised by two Master of Public Health professionals.

### Data Quality Assurance

The English version of the questionnaire was translated to local language Afaan Oromo and then translated back to English by two fluent speakers in order to check consistency. The questionnaire was pretested in 5% of the non-study population at Bilo Karo health center, and some adjustments were made to the questionnaire accordingly. Data collectors and supervisors were given two days training about the objective and purpose of the study including techniques of data collection. The completeness and clarity of questionnaires were checked after data collection. Principal investigators checked questionnaires visually for incompleteness and appropriate measures were taken before the next data collection day. The possible effect of a confounder was controlled through multivariate logistic regression.

### Data Processing and Analysis

The collected data were coded and entered into Epi data 3.1 version statistical packages and exported to SPSS version 24 for analysis. Both descriptive and analytical statistics were used. Descriptive result was presented using tables and figure. Bivariate logistic regression analysis was used to identify associations between variables. Then, variables whose p<0.2 in the bivariate analysis were taken as candidate for multivariate analysis. Multivariate logistic regression analysis was used to identify the predictors of the outcome variable. The fitness of the model was tested by Hosmer-Lemeshow goodness of fit test. Odd Ratios along with 95% Confidence interval was calculated to measure the strength of the association. Association between the explanatory and dependent variable was assessed at a p-value of 0.05.

### Ethical Consideration

Ethical clearance was received from Ethical Review Committee of Mettu University. Confidentiality was assured by excluding personal identifiers such as name. The study purpose, procedure and duration, possible risks and benefits of the study were clearly explained for study participants and written informed consent was obtained from respondents.

## Results

### Socio-Demographic and Professional Characteristics

Two hundred forty-one obstetrics care givers were participated in the study, yielding a response rate of 96.4%. The mean age of the participants was 28.86 years (standard deviation= ±3.48 years). Most (56.8%) of obstetric care givers were females. Almost half (50.6%) of the obstetrics care givers had more than five years of service. According to respondents, almost all (94.6%) of them had attended less than 10 deliveries per a day, and majority (92.5%) of them said that there were less than four birth attendants per a working day (Table1).

**Table 1:** Socio-demographic and professional characteristics among obstetric care givers in Ilu Aba Bor Zone, southwest Ethiopia, 2020 (n = 241).

| Variables | Category | Frequency |  |
| --- | --- | --- | --- |
|  |  | Number | Percent |
| Sex of respondent | Male | 104 | 43.2 |
|  | Female | 137 | 56.8 |
| Educational status | Diploma | 96 | 39.8 |
|  | BSc (Bachelor of Science) and above | 145 | 60.2 |
| Work place | Health center | 207 | 85.9 |
|  | Hospital | 34 | 14.1 |
| Profession | Midwifery | 89 | 36.9 |
|  | Other | 152 | 63.1 |
| Service year | <2 | 40 | 16.6 |
|  | 2-5 | 79 | 32.8 |
|  | >5 | 122 | 50.6 |
| Regularly working department | Delivery | 35 | 14.5 |
|  | Other | 206 | 85.5 |
| Learned about partograph academically | No | 32 | 13.3 |
|  | Yes | 209 | 86.7 |
| On-the-job training on partograph received | No | 130 | 53.9 |
|  | Yes | 111 | 46.1 |
| Number of deliveries per a day | <10 | 228 | 94.6 |
|  | ≥10 | 13 | 5.4 |
| Number of birth attendants per a working day | <4 | 223 | 92.5 |
|  | ≥4 | 18 | 7.5 |
| Partograph available | No | 106 | 44.0 |
|  | Yes | 135 | 56.0 |

### Utilization of Partograph

Although 181 (75.1%) of all obstetric care givers used partograph in labor management, only 79 (32.8%) of all obstetric care givers used partograph routinely. Regarding recording labor activities information on partograph, 65.1%, 57.3% and 42.7% of obstetric care givers had recorded the information on partograph about cervical dilatation every 4 hours, fetal heart beat every 30 minutes, and maternal pulse rate every 30 minutes, respectively (Table 2).

**Table 2:** Utilization of partograph among obstetric care givers in Ilu Aba Bor Zone, southwest Ethiopia, 2020 (n = 241).

| Variables | Category | Frequency |  |
| --- | --- | --- | --- |
|  |  | Number | Percent |
|  |  | (n) | (%) |
| Use partograph in labor management | No | 60 | 24.9 |
|  | Yes | 181 | 75.1 |
| Partograph use frequency | Routinely | 79 | 32.8 |
|  | Sometimes | 102 | 42.3 |
| Uterine contraction plotted half hourly | No | 68 | 28.2 |
|  | Yes | 113 | 46.9 |
| Initial cervical dilatation plotted | No | 22 | 9.1 |
|  | Yes | 159 | 66.0 |
| Membrane intactness recorded | No | 68 | 28.2 |
|  | Yes | 113 | 46.9 |
| Cervical dilatation plotted every 4hr | No | 24 | 10.0 |
|  | Yes | 157 | 65.1 |
| Descent plotted every 4hr | No | 68 | 28.2 |
|  | Yes | 113 | 46.9 |
| Fetal heart beat recorded every 30 min | No | 43 | 17.8 |
|  | Yes | 138 | 57.3 |
| Color of liquor recorded | No | 68 | 28.2 |
|  | Yes | 113 | 46.9 |
| Maternal blood pressure monitored every 4 hours | No | 69 | 28.6 |
|  | Yes | 112 | 46.5 |
| Maternal pulse rate monitored every 30 minutes | No | 78 | 32.4 |
|  | Yes | 103 | 42.7 |
| Action taken based on partograph | No | 6 | 2.5 |
|  | Yes | 175 | 72.6 |

Of the 60 obstetric care givers that do not use partograph, the top three reasons for not utilizing partograph were lack of partograph (56.7%), poor managerial support (45.0%) and absence of training of health professionals (33.3%) (Fig 1).

**Fig 1:**
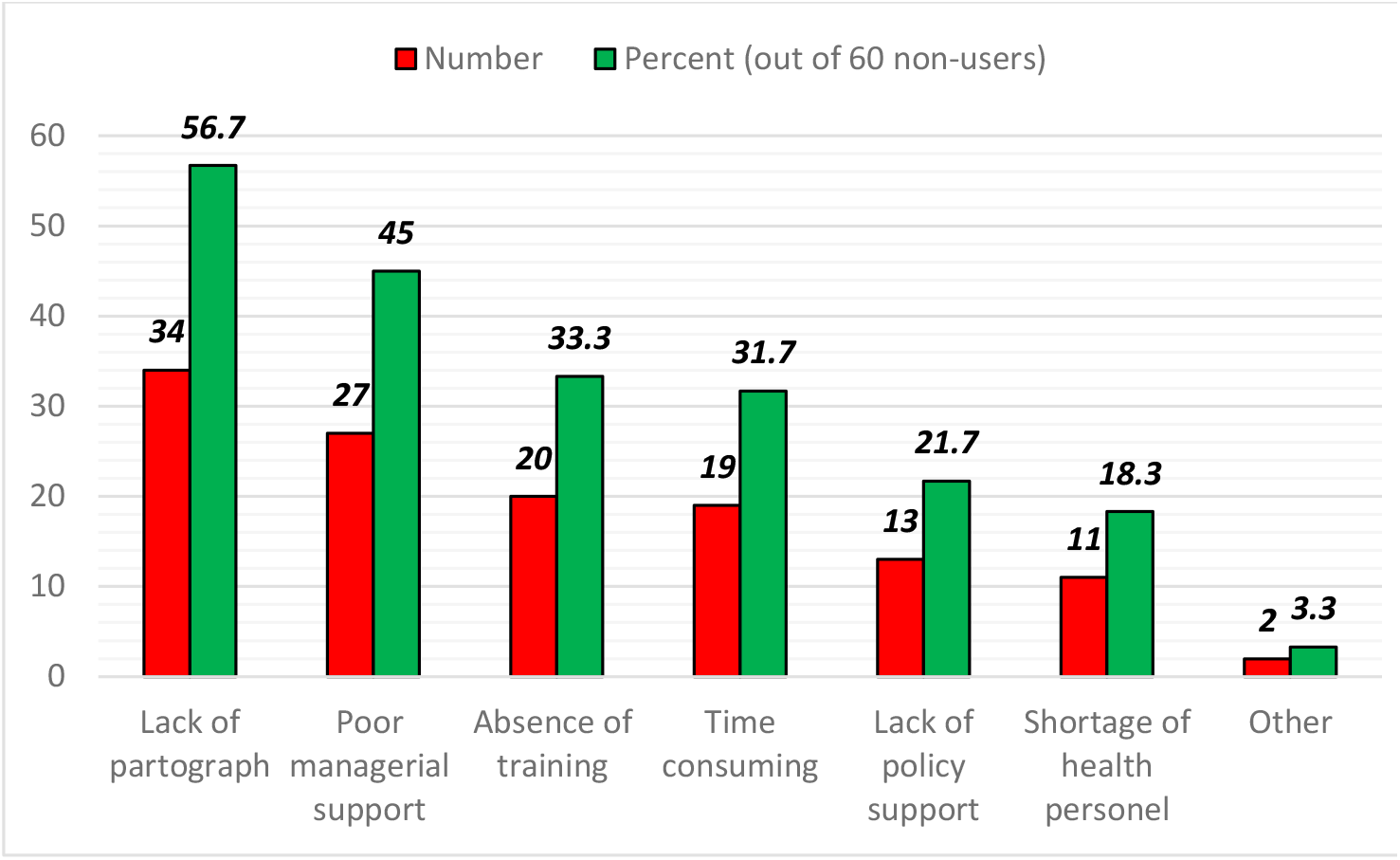
Reasons for not using partograph among obstetric care givers in Ilu Aba Bor Zone, southwest Ethiopia, 2020 (n = 60).

### Factors Associated with Partograph Utilization among Obstetric Caregivers

Using bivariate logistic regression analysis, the following ten variables were associated with partograph utilization: service year, on-job training on partograph, work place, educational status, regularly working department, profession, partograph availability, learning about partograph academically, number of deliveries per a day, and number of birth attendants per a day. However, only four variables (on-job training on partograph, work place, educational status, and partograph availability) were significantly associated with partograph utilization during multivariable analysis at p-value less than 0.05 with Hosmer and Lemeshow goodness of fit p=0.867 (Table 3).

**Table 3:**
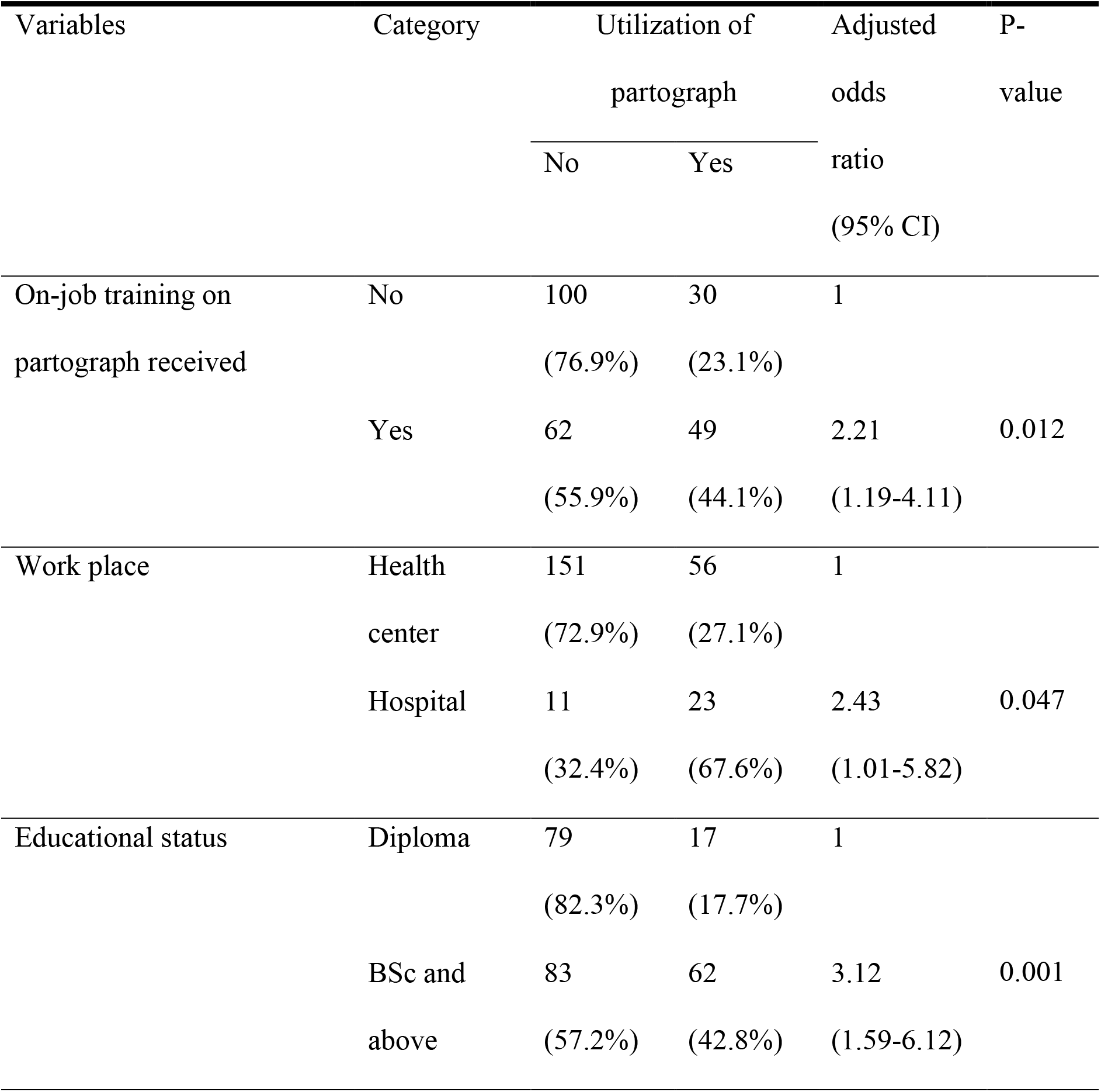
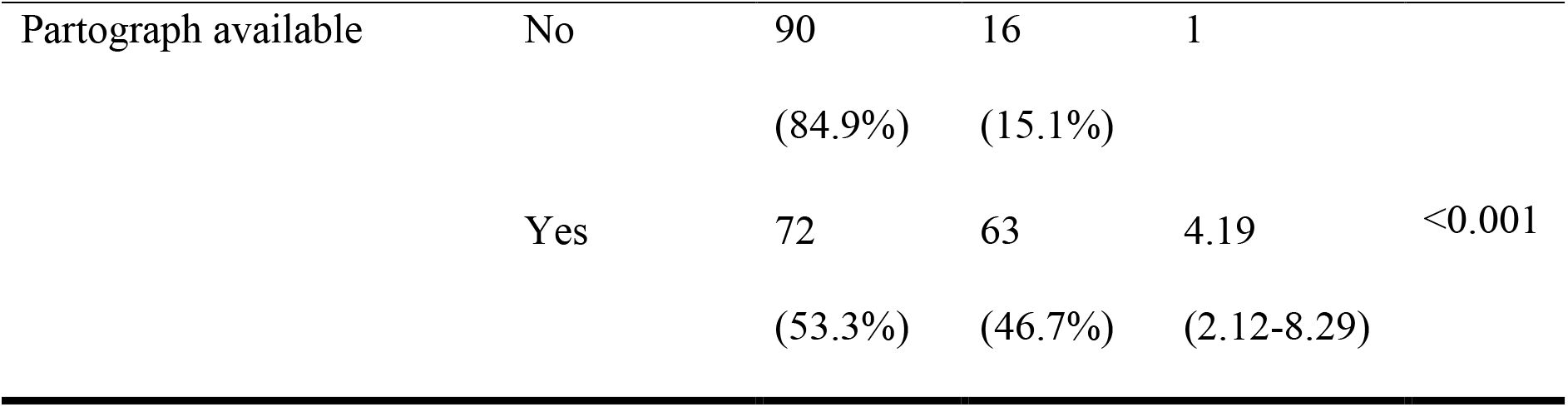
Multivariable analyses for factors associated with partograph utilization among obstetric care givers in Ilu Aba Bor Zone, southwest Ethiopia, 2020 (n = 241).

This study found that those obstetric caregivers who received on-job training on partograph were almost two times more likely to use partograph compared to those who did not receive on-job training (Adjusted odds Ratio (AOR) = 2.21, 95% Confidence Interval (CI)= 1.19, 4.11). Obstetric care givers who were working in a hospital were more likely to use partograph than those who were working in a health center (AOR = 2.43, 95%CI = 1.01, 5.82), while having BSc and above educational status had tripled likelihood of using partograph in contrast to having Diploma educational status (AOR = 3.12, 95%CI = 1.59, 6.12). The study revealed that those participants who had partograph in their health facility were almost four times more likely to utilize partograph compared to those who had no partograph (AOR = 4.19, 95%CI = 2.12, 8.29). Variables Category Utilization of partograph

## Discussion

This study involved obstetrics care givers working in selected public health institution of Ilu Aba Bor Zone to determine level of partograph utilization and associated factors. Eventually, it was found that only one third (32.8%) of participants were using partograph routinely as WHO recommendation, and on-job training on partograph, work place, educational status, and partograph availability were factors associated with partograph utilization.

The level of partograph utilization routinely in this study is lower than those studies conducted in Tigray Region, Northern Ethiopia (73.3%),^23^ and in the Eastern Province of Rwanda (41.22%).^26^ The reason for higher partograph utilization in Tigray Region might be because of more participants with in service training about partograph. Similarly, more obstetrics caregivers in the study done in Eastern Province of Rwanda received training on partograph than this study, which obviously increase knowledge about partograph that in turn boost the tendency to use the chart. Moreover, difference in study area and population might be the reason for the discrepancy.

Lack of partograph, poor managerial support, absence of training of health professionals, being time consuming, lack of policy support, and shortage of health personnel were reasons for not utilizing partograph among obstetrics caregivers that were not utilizing the partograph. These reasons were in agreement with other studies conducted in Ethiopia and South Africa.^12,24,28^

This study revealed that those obstetric caregivers who received on-job training on partograph were almost two times more likely to use partograph compared to those who did not receive on-job training. This finding was in line with other studies conducted in the Ethiopia, Kenya and Rwanda.^11,21,26^ The association between on-job training on partograph and use of partograph might be because training increase knowledge and attitude of participants about partograph that could in turn increase partograph use.

Obstetric care givers who were working in a hospital were more likely to use partograph than those who were working in a health center. This association was also observed in the study done in Hadiya Zone, Southern Ethiopia.^19^ This could be because of the higher chance of access to information, infrastructures and training among obstetrics caregivers that works in hospital, that are usually located in urban area, compared to participants who were working in a health center.

Those obstetrics caregivers with BSc and above educational status were three times more likely to use partograph in contrast to those with Diploma educational status. This finding is in agreement with studies carried out in East Gojam Zone, Northern Ethiopia.^24^ This association is might be due to the fact that participants with BSc and above educational status have more comprehensive and elaborated knowledge about partograph compared to those with Diploma, and that will increase their possibility of using partograph.

Furthermore, this study revealed that those participants who had partograph in their health facility were almost four times more likely to utilize partograph compared to those who had no partograph. The finding is in line with studies conducted in Ethiopia, as well as in Cross River State and the Niger Delta Region of Nigeria.^20,29^ This might be because a higher availability of partograph in the health facilities of obstetrics caregivers increases exposure and willingness of partograph use.

Due to the employment of cross-sectional study design, this study has weakness of not showing the temporal cause effect relationship between the partograph use and associated factors. Since self-administered questionnaires were used, there could be possibility of social desirability bias. Moreover, it could have been better to include obstetrics caregivers of private institutions in the study.

## Conclusion and Recommendations

This study found out that partograph utilization among obstetric care givers working in public health institutions found in Ilu Aba Bor Zone was very low. On-job training on partograph, work place, educational status, and partograph availability were factors associated with partograph utilization among the obstetric care givers. Therefore, the authors recommend concerned bodies to strive in order to achieve WHO recommendation of routine partograph use among obstetric care givers. Accordingly, efforts should be exerted to make partograph and infrastructures available in health facilities. Moreover, providing on-job training and academic development opportunity to obstetric care givers is anticipated to increase partograph utilization.

## Data Availability

All data used for the current study are presented in the manuscript.

## Acknowledgments

The authors would like to acknowledge staffs of Mettu University Public Health Department for their constructive comments and ideas. The researchers are highly thankful to Ilu Aba Bor Zone Health Department for providing some important information. The authors would like to extend their appreciation to managers and participants of public health institutions in Ilu Aba Bor Zone for their great cooperation.

## Abbreviations

AOR: Adjusted odds ratio
BSc: Bachelor of Science
CI: Confidence Interval
WHO: World Health Organization

## Authors & contributions

Fentaneh Teshome and Ebissa Negera conceived the study, developed the tool, coordinated data collection, carried out the statistical analysis, and drafted the manuscript. Getamesay Aynalem was involved in designing the study, data analysis and interpretation. All authors read and approved the final manuscript.

## Data Availability

All data used for the current study are presented in the manuscript.

## References

1. World Health Organization, “Trends in maternal mortality: 1990 to 2015: estimates by WHO, UNICEF, UNFPA,” World Bank Group and the United Nations Population Division. Geneva, Switzerland, 2015.

2. Central Statistical Agency (CSA) [Ethiopia] and ICF. 2016. “Ethiopia Demographic and Health Survey 2016: Key Indicators Report.” Addis Ababa, Ethiopia, and Rockville, Maryland, USA. CSA and ICF.

3. World Health Organization, “Reduction of maternal mortality: a joint WHO/ UNFPA/ UNICEF/ World Bank statement,” Geneva, Switzerland, 1999.

4. World Health Organization, “Mother-baby package: Implementing safe motherhood in countries,” Geneva, Switzerland, 1994.

5. World Health Organization, “The partograph: the application of the WHO partograph in the management of labor,” Geneva, Switzerland, 1994.

6. M. Mathai, “The Partograph for the Prevention of Obstructed Labor,” Clinical Obstetrics and Gynecology, vol. 52, no. 2, pp. 256–269, 2009.

7. G. Dangal, “Preventing prolonged labor by using partograph,” The Internet Journal of Gynecology and Obstetrics, vol. 7, no. 1, 2007.

8. S. Tayade and P. Jadhao, “The impact of use of modified who partograph on maternal and perinatal outcome,” International Journal of Biomedical and Advance Research, vol. 3, no. 4, 2012.

9. B. Ahmed, et al., “Partograph versus no partograph: effect on labor progress and delivery outcome: a comparative study,” International Journal of Reproduction, Contraception, Obstetrics and Gynecology, vol. 6, no. 11, pp. 4928–4933, 2017. http://dx.doi.org/10.18203/2320-1770.ijrcog20175002

10. E. Ollerhead and D. Osrin, “Barriers to and incentives for achieving partograph use in obstetric practice in low- and middle-income countries: a systematic review,” BMC Pregnancy and Childbirth, vol. 14, no. 281, 2014. http://www.biomedcentral.com/1471-2393/14/281

11. C.N. Githae, A. Mbisi and J.O. Boraya, “Utilization of Partograph in the Management of Women in Labor among Nurses Working in Embu County, Kenya,” Nursing Healthcare International Journal, vol. 3, no. 2: 000181, 2019. DOI: 10.23880/nhij-16000181

12. O.M. Maphasha, I. Govender, D.P. Motloba and C. Barua, “Use of the partogram by doctors and midwives at Odi District Hospital, Gauteng, South Africa,” South African Family Practice, vol. 1, no. 1, pp. 1–5, 2017. http://dx.doi.org/10.1080/20786190.2017.1280899

13. B.K. Opoku and S.B. Nguah, “Utilization of the modified WHO partograph in assessing the progress of labor in a metropolitan area in Ghana,” Research Journal of Women ‘s Health, vol. 2, no. 2, 2015. http://dx.doi.org/10.7243/2054-9865-2-2

14. S. Ogwang, Z. Karyabakabo and E. Rutebemberwa, “Assessment of partogram use during labor in Rujumbura Health Sub District, Rukungiri District, Uganda,” African Health Sciences, vol. 9, no. S2, pp. 27–34, 2009.

15. M.M. Opiah, A.B. Ofi, E.J. Essien and E. Monjok, “Knowledge and Utilization of the Partograph among Midwives in the Niger Delta Region of Nigeria,” African Journal of Reproductive Health March, vol. 16, no. 1, pp. 125–132, 2012.

16. Federal Democratic Republic of Ethiopia, “Ethiopia 2017 Voluntary National Review on SDGs: Government Commitments, National Ownership and Performance Trends,” National Planning Commission, Addis Ababa, Ethiopia, 2017.

17. A. Getachew, J. Ricca, D. Cantor, et al., “Quality of Care for Prevention and Management of Common Maternal and Newborn Complications: A Study of Ethiopia ‘s Hospitals,” Jhpiego, Maryland, USA, 2011.

18. D. Bekele, K. Beyene, L. Hinkosa and M. N. Shemsu, “Partograph Utilization and associated Factors among Graduating Health Professional Students in Asella Referal and Teaching Hospital, Ethiopia, 2016,” Research Reviews: A Journal of Computational Biology, vol. 6, no. 2, pp. 12–18, 2017.

19. Y. Haile, F. Tafese, T. D. Weldemarium and M. H. Rad, “Partograph Utilization and Associated Factors among Obstetric Care Providers at Public Health Facilities in Hadiya Zone, Southern Ethiopia,” Hindawi Journal of Pregnancy, vol 2020, Article ID 3943498, 8 pages, 2020. https://doi.org/10.1155/2020/3943498

20. U. Asibong, I.B. Okokon, T.U. Agan, et al., “The use of the partograph in labor monitoring: a cross-sectional study among obstetric caregivers in General Hospital, Calabar, Cross River State, Nigeria,” International Journal of Women ‘s Health, vol. 2014, no. 6, pp. 873–880, 2014. http://dx.doi.org/10.2147/IJWH.S49188

21. N. Wakgari, A. Amano, M. Berta and, G.A. Tessema, “Partograph utilization and associated factors among obstetric care providers in North Shoa Zone, Central Ethiopia: a cross sectional study,” African Health Sciences, vol. 15, no. 2, 2015. http://dx.doi.org/10.4314/ahs.v15i2.30

22. M. Markos, A. Arba and K. Paulos, “Partograph Utilization and Associated Factors among Obstetric Care Providers Working in Public Health Facilities of Wolaita Zone, 2017,” Hindawi Journal of Pregnancy, vol 2020, Article ID 3631808, 8 pages, 2020. https://doi.org/10.1155/2020/3631808

23. T. Hailu, K. Nigus, G. Gidey, B. Hailu and Y. Moges, “Assessment of partograph utilization and associated factors among obstetric care givers at public health institutions in central Zone, Tigray, Ethiopia,” BMC Research Notes, vol. 11, no. 710, 2018. https://doi.org/10.1186/s13104-018-3814-7

24. D.A. Zelellw and T.K. Tegegne, Level of partograph utilization and its associated factors among obstetric caregivers at public health facilities in East Gojam Zone, Northwest Ethiopia,” PLoS ONE, vol. 13, no. 7: e0200479, 2018. https://doi.org/10.1371/journal.pone.0200479

25. H. Regasa, T. Tilahun and H. Adem, “Utilization of partograph and associated factors among obstetric care givers in hospitals of Western Oromia, Ethiopia, 2017,” Panacea Journal of Medical Sciences, vol. 8, no. 1, pp. 21–24, 2018. DOI: 10.18231/2348-7682.2018.0005

26. O. Bazirete, N. Mbombo and O. Adejumo, “Utilisation of the partogram among nurses and midwives in selected health facilities in the Eastern Province of Rwanda,” Curationis, vol. 40, no. 1, pp. a1751, 2017. https://doi.org/10.4102/curationis.v40i1.1751

27. T.O. Egbe, E.N Ncham, W. Takang, E-N Egbe and G.E. Halle-Ekane, “Use of the partogram in the Bamenda health district, north-west region, Cameroon: a cross-sectional study,” Gynecology and Obstetrics Research Open Journal, vol. 2, no. 5, pp. 102–111, 2016. http://dx.doi.org/10.17140/GOROJ-2-124

28. E. Yisma, B. Dessalegn, A. Astatkie and N. Fesseha, “Completion of the modified World Health Organization (WHO) partograph during labor in public health institutions of Addis Ababa, Ethiopia,” Reproductive Health, vol. 10, no. 23, 2013. http://www.reproductive-health-journal.com/content/10/1/23

29. W. Willi and M. Molla, “Partograph chart use among obstetric caregivers in public health institutions of West Shewa Zone, Oromia Regional State, Ethiopia, 2015,” Ethiopian Journal of Reproductive Health, vol. 9, no. 1, pp. 36–44, 2017.

